# PARASITIC CONTAMINATION OF RAW FRESH FRUITS AND VEGETABLES COMMONLY SOLD AT CENTRAL MARKETS IN BAMENDA MUNICIPALITY, NORTH WEST REGION - CAMEROON

**DOI:** 10.1101/2025.05.22.25328157

**Authors:** Nguemaïm N Flore, Nyong H Ndukong, Farikou Oumarou, Kamga HL Fouamno

**Author notes:** Corresponding author: Nguemaïm Ngoufo Flore.

## Abstract

Consumption of raw fruits and vegetables constitute a potential source of spread for various parasitic infections. The level of contamination and the various species of parasites found are based on climatic, ecological, and human factors. The purpose of this study was to determine local data about the contamination status of raw fruits and vegetables commonly sold in markets at the Bamenda municipality and the predisposing factors, for better control of parasitic diseases. A cross-sectional study was conducted from May to July 2023 on fruits and vegetables collected from local markets of Bamenda town. A total of 200 samples were purchased from four main markets located at center town. The samples were microscopically examined for detection of different parasitic forms and determine the level of parasitic contamination using standardized parasitological techniques for protozoans and helminthes. The difference between prevalence of intestinal parasites among different categories was compared using Pearson chi-square test. Univariate logistic regression was used to assess factors associated with parasitic contamination of fruits and vegetables. The result was considered statistically significant when *p* value was less than 0.05 at 95% confidence level. A total of 123 individual parasites were found representing 61.5% of the infection in total produce. Ten (10) protozoa parasites were detected and *Cryptosporidium spp* (25.0%) were more represented followed by *Entamoeba histolytica* (12.5%) while *Entamoeba coli* (2.0%), *Isospora belli* (2.0%) and *Balantidium coli* (2.0%) have the least rate. Six (6) species of helminth parasites were identified and Hookworms (3,5%) were more present. The overall prevalence of parasitic contamination was 47.0%. Washing of the fruits and vegetables before display for selling was significantly associated with decreased parasitic contamination (*p* < 0.05). The overall prevalence of parasitic contamination was 47.0%. Different parasites were identified in mono and or in polyinfection. *C. pavum* and *E. histolytica* were protozoa more common, while *A. lumbricoïdes* and Hookworms were the most frequent helminths. The act of washing fruits and vegetables before displaying for sale, the sources of water used for washing, the means of display for selling and their duration in the market appeared to be factors associated with parasitic contamination.

## INTRODUCTION

Food-borne illnesses caused by intestinal parasites are still a public health problem in developing countries [1]. More than two billion of people in the world are infected with intestinal parasites [2]. Fresh vegetables and fruits are an important source of nutrients and bioactive non-nutrient plant compounds including vitamins, minerals, phytochemicals especially antioxidants, and dietary fibers [3, 4]. Daily use of fruits and vegetables can reduce the risk of chronic diseases (diabetes, cardiovascular disease, cancer, age-related diseases) and improve the function of the digestive system. Thus, nutritionists recommend eating fresh fruits and vegetables in daily diet [5]. In order to preserve the flavor and nutrients of vegetables, they are usually consumed raw, which can increase the risk of infectious parasitic diseases [6, 7]. Eating unclean, raw, or undercooked fruits and vegetables is the most important way of transmission of intestinal parasites which is often considered as a major concern of public health. Protozoan cysts, worm eggs, larvae usually survive and develop well in moist soil and environments of farm vegetables necessary for continuation of soil-transmitted parasite life cycle [8 - 10].

Fruits and vegetables act as vehicles for the transmission of parasitic infections when contaminated as a result of various associated factors related to planting, such as while they are still on the field, harvesting, transportation, storage, market chain, and even at home [11, 12]. The cultivation of vegetables in many parts of the world has been amplified with the application of fertilizer and or manure. In Africa, the transmission of intestinal parasitic infection has been considered to increase successfully due to the frequent use of untreated human or animal dung as manure in cultivation by the local farmers, which serves as a source of enhancement of zoonotic parasitic infection [13].

Ingestion of viable infective protozoan cysts occurs in amoebiasis caused *Entamoeba histolytica*, in giardiasis caused by *Giardia lamblia* and in infections caused by *Isospora belli, Cryptosporidium parvum, Sarcosystis hominis* and the ciliate *Balantidium coli*.

Furthermore, intestinal infection by *Trichomonas hominis* involves the ingestion of viable trophozoites, which serve as the infective stage. Ingestion of viable, infective embryonated eggs eaten with contaminated food or drink occurs in ascariasis caused by *Ascaris lumbricoides* [14], Trichuriasis caused by *Trichuris triuchiura* and occasionally, enterobiasis caused by *Enterobius vermicularis*, hymenolepiasis caused by *Hymenolepis nana*, and cysticercosis caused by *Taenia solium tapeworms*. Moreover, consumption of raw or unhygienically prepared vegetables such as cabbage (*Brassica oleracea*), garden egg (*Solanum melongena*), cucumber (*Cucumis sativus*), lettuce (*Lactuca sativa*), carrot (*Daurus carota*), waterleaf (*Talinum trangulare*), pumpkin (*Cucurbita pepo*), tomatoes (*Solanum lycopersicum*), etc, is considered a risk factor for human parasitic infections [13 - 14]. Studies have shown that *Ascaris lumbricoides, Cryptosporidium* spp., *Entamoeba histolytica, Enterobius vermicularis, Fasciola* spp., *Giardia intestinalis, hookworm, Hymenolepis* spp., *Taenia* spp., *Trichuris trichiura, and Toxocara* spp., can infect humans who consume contaminated, uncooked, or improperly washed vegetables and fruits [15]. Studies of Kozan *et al*. [16], Said [17] and Hassan [18] conducted to evaluate the role of raw vegetables in the transmission of intestinal parasites have stressed the importance of fruits and vegetables, particularly when they are consumed raw and unwashed, in the transmission of medically important parasites.

According to Abougrain *et al*.[19], there is a strong association between raw fruits and vegetables, and parasitic infections. Moreover, many outbreaks of protozoan infections in humans have been linked to raw fruits and vegetables [20]. Intestinal parasitic infections are widely distributed throughout the world causing substantial intimidation to the public health, economy, and physical and cognitive development particularly among children in developing countries. The poor personal hygiene, poor environmental hygiene, and poor health system commonly observed make the prevalence to be highest among the populations [21,22].

In Cameroon, poor sanitation, and substandard of living conditions lead to an increased risk of acquiring parasitic infections. Despite the fact that human intestinal parasitosis is common in Bamenda [23, 24]. Data on parasitic contamination of fruits and vegetables, sold in markets within the city are very scarce. This study aims to determine local data about the contamination status of raw fruits and vegetables commonly sold in markets at the Bamenda municipality and the predisposing factors, for better control of parasitic diseases.

## MATERIAL AND METHODS

### Study design and study area

A cross-sectional study was conducted from May to July 2023 on fruits and vegetables collected from local markets of Bamenda town. The city is located in the North West region of Cameroon. It has a population of about 2 million people and situated at 366 kilometres from the capital Yaoundé. It is located at 5°56’ north latitude and 10°10’ east longitude. The main industries are the processing of agricultural produce such as elementary food. The weather condition in and around the city is conducive for cultivation of diverse items of fruits and which are readily available in local markets throughout the town and most items are eaten raw. Main market, Food market, Ntarinkon market and Nkwen market are four randomly selected markets of the center town where the majority of fruits and vegetables are brought directly from farmers or middlemen in order to sell to consumers. Data for the present study was collected from those markets.

### Data collection

A structured questionnaire was self administered to vendors to collect data on sociodemographic characteristic of vendor as well as health related factor through face to face interview and observation.

A total of 200 samples of fruits and vegetables were purchased from the markets. The samples were made of five fruit types: pineapple (*Ananas comosus*), watermelon (*Citrullus lanatus*), orange (*Citrus sinensis*), mango (*Mangifera indica*) and pawpaw (*Carica papaya*) and five vegetable types: Cucumber (*Cucumis sativus*), cabbage (*Brassica oleracea*), carrot (*Daucus carota*), tomato (*Solanum lycopersicum*) and garden egg (*Solanum melongena*). Equal numbers of samples (20 each) were bought from the selected markets (50 samples per market) and put in a sterile plastic bag properly labeled, and transported in a cold box to the Medical Parasitology Department of the Bamenda Regional Hospital Laboratory for parasitological examination.

### Laboratory procedure

A portion of 100 g of each fruit and vegetable was washed separately in a beaker containing 250 mL of normal saline (0.85% NaCl) for 20 min followed by agitation in a shaker for 5min for adequate washing [25, 26]. The sample were then remove and the washing solution was incubated overnight for adequate sedimentation of ova, larvae and cysts of helminths and protozoan parasites assumed associated with fruit and /or vegetable contamination. After overnight sedimentation of the washing solution, 5mL of the sediment was then transferred to a centrifuge tube using a sieve to remove undesirable particles. The tube content was centrifuged at 3000 rpm for 5 min in order to concentrate the parasitic stages [27]. After centrifugation, the supernatant was decanted carefully without shaking and the sediment was agitated gently by hand for resuspend the sediment. A part of it was then picked using a Pasteur pipette to prepare direct wet mount smears for microscopic observations. The preparation was examined under light microscope for detection of parasite stages using the x10 objective lens. Definite morphological features were identified under the x40 objective. The remaining sediment was used to prepared smears by Modified Ziehl-Neelsen staining techniques for the detection of coccidian protozoan oocysts including *Cryptosporidium spp*., *Isospora belli*, and *Cyclospora cayetanensis* following the protocol explained by Tefera *et al*. [28] and Ebrahimzadeh *et al*. [10]. These slides were observed using the x100 objective. Slides were prepared and examined per sample both in direct wet mount and Modified Ziehl-Neelsen staining techniques.

### Data management and statistical analysis

Data were entered in Microsoft excel 2016 and analyzed using SPSS version 20. Descriptive analysis such as frequency and proportion were calculated to explain characteristics of vendor and the level of contamination of fruits and vegetables collected. The difference between prevalence of intestinal parasites among different categories was compared using Pearson Chi-square test. Univariate logistic regression was used to assess factors associated with parasitic contamination of fruits and vegetables. The result was considered statistically significant when *p* value was less than 0.05 at 95% confidence level. Results were presented in the form of tables.

### Ethical considerations

Ethical clearance was obtained from Institutional Review Board of the University of Bamenda located in the Faculty of Health Science. Administrative approvals were obtained from the General supervisor of the Bamenda Regional Hospital and the head of the Parasitology unit of the Laboratory where the research was carried out. Signed inform consent was obtained from each vendor.

## RESULTS

### General characteristics of study participants

A total of 200 vendors were enrolled in this study, 190 were females (95.0%) and 10 (5.0%) were males. Participants between 20 to 35 years the most represented age group (45.5%) while those above the age of 50 years were the least represented (6.5%). With regards to the educational status, the majority of the vendors (53.0%) had secondary education, followed by 40.0% who have attained primary education, while 6.0% and 1% had respectively tertiary and no formal education (Table 1).

**Table 1:** Sociodemographic characteristics of study participants (N = 200)

| Variables | Frequency | Percentage |
| --- | --- | --- |
| **Gender** |  |  |
| Male | 10 | 5 |
| Female | 190 | 95 |
| **Age ranges (years)** |  |  |
| <20 | 28 | 14.0 |
| 20-35 | 91 | 45.5 |
| 36-50 | 68 | 34.0 |
| >50 | 13 | 6.5 |
| **Level of education** |  |  |
| No formal education | 2 | 1.0 |
| Primary | 80 | 40.0 |
| Secondary | 106 | 53 |
| Tertiary | 12 | 6.0 |

N: total number

### Parasites contamination rate of fruits and vegetables collected

A total of 200 samples of fruits and vegetables were collected from 200 vendors of 4 local markets and examined for parasitological contamination. Table 2 showed that 47.0% of samples were identified to be contaminated with at least one type of parasite. Carrot appeared to be the most frequently contaminated (70.0%) among all the items while pawpaw (35.0%) had the least parasite contamination. The result indicated that vegetables were more contaminated than fruits that are 51% and 43% respectively.

**Table 2:** Distribution of parasitic contamination among fruits and vegetables collected.

| Different products |  | Number examined | Number positive (%) | Number of parasite detected (%) |  |  | Number negative (%) |
| --- | --- | --- | --- | --- | --- | --- | --- |
|  |  |  |  | One | Two | Three and above |  |
| **Fruits** | Mango | 20 | 07 (35.0) | 02(10.0) | 04(20.0) | 01(5.0) | 13(65.0) |
|  | Pawpaw | 20 | 05(25.0) | 04(20.0) | 01(5.0) | 00 | 15(75.0) |
|  | Orange | 20 | 06 (30.0) | 05(25.0) | 01(5.0) | 00 | 14(70.0) |
|  | Watermelon | 20 | 13 (65.0) | 08(40.0) | 04(20.0) | 01(5.0) | 7(35.0) |
|  | Pineapple | 20 | 12 (60.0) | 07(35.0) | 04(20.0) | 01(5.0) | 8(40.0) |
| **Total 1** |  | **100** | **43(43.0)** | **26(26.0)** | **14(14.0)** | **3(3.0)** | **57(57.0)** |
| **Vegetables** | Garden egg | 20 | 08 (40.0) | 07(35.0) | 01(5.0) | 00 | 12(60.0) |
|  | Carrot | 20 | 14 (70.0) | 09(50.0) | 05(25.0) | 00 | 6(30.0) |
|  | Cucumber | 20 | 08 (40.0) | 05(25.0) | 03(15.0) | 00 | 12(60.0) |
|  | Tomato | 20 | 08 (40.0) | 05(25.0) | 03(15.0) | 00 | 12(60.0) |
|  | Cabbage | 20 | 13 (65.0) | 07(35.0) | 06(30.0) | 00 | 7(35.0) |
| **Total 2** |  | **100** | **51(51.0)** | **33(33.0)** | **18(18.0)** | **00** | **49(49.0)** |
| **Total 1+2** |  | **200** | **94 (47.0)** | **59 (29.5)** | **32(16.0)** | **03(1.5)** | **106(53.0)** |

Polyparasitic contamination was observed both in fruits and vegetables. It appeared that 29.5% of the total sample was contaminated with one type of parasite. Biparasitic contamination was found in 16.0% while 1.5% of the samples had at least three species of parasites (noticed in fruits only). For fruits, watermelon appeared more contaminated (65.0%), followed by pineapple (60.0%), mango (35.0%), orange (30.0%), and then pawpaw (25.0%). For vegetables, carrot were more contaminated (70.0%) followed by cabbage (65.0%) and the rest (garden egg, cucumber, tomato) had equal proportion of parasites that is 40% each.

The parasites encountered include species of protozoa and helminths. A total of 123 individual parasites were found representing 61.5% of the infection in total produce. Ten (10) protozoa parasites were detected and *Cryptosporidium spp* (25.0%) were more represented followed by *Entamoeba histolytica* (12.5%) while *Entamoeba coli* (2.0%), *Isospora belli* (2.0%) and *Balantidium coli* (2.0%) have the least rate. Six (6) species of helminth parasites were identified and Hookworms (3,5%) were more present. Unidentified parasite representing a rate of 1.6% was found. Helminths were not found both on mango and cucumber (Table 3).

**Table 3:**
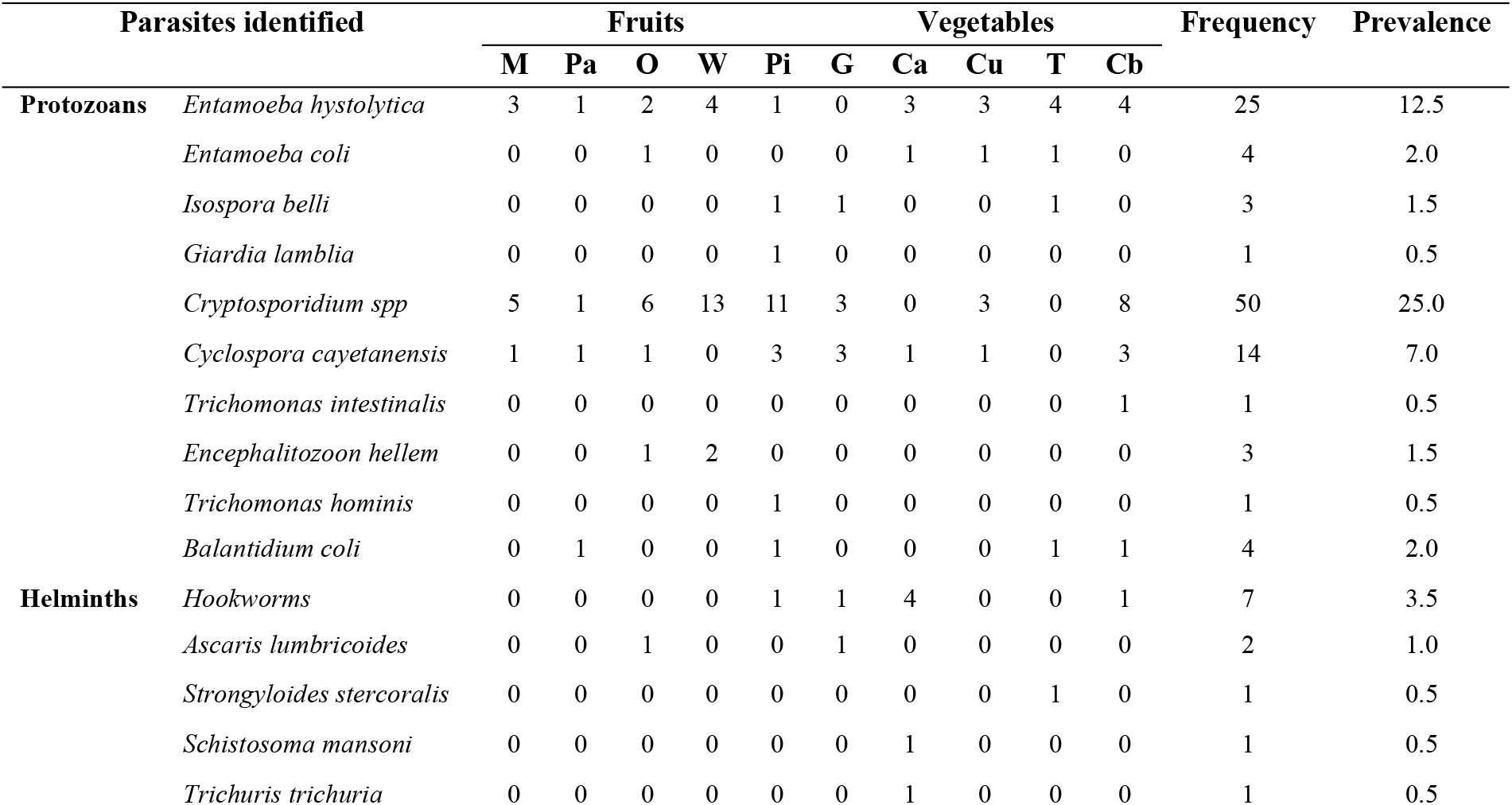
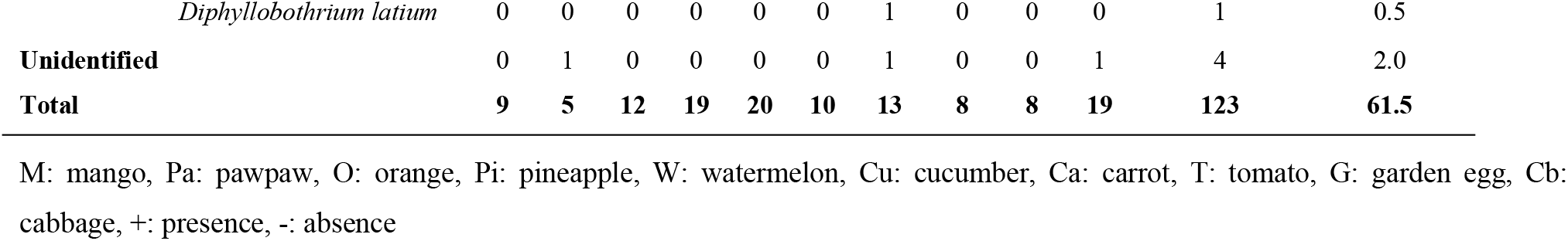
Frequency and prevalence of intestinal parasites found in fruits and vegetables (N = 200)

### Factors associated with parasite contamination of fruit and vegetables collected

Amongst the factors associated with contamination, produce of the female vendor were more contaminated. It was the same with those aged between 36 to 50 years of age. Vendor with no formal education and those with primary education were found with produce more contaminated. The difference between the rates of contamination among the different levels of education of vendor was statistically significant (*p* < 0.05). More than 50% produce contaminated were found in vendor with untrimmed fingernail.

The results showed that the difference between the means of display of produce was statistically significant (*p* < 0.05). The sample collected from Main market had high contamination rate (28.0%) followed by samples collected from Ntarinkon market (26.9%), Foot market (25.8%), and Nkwen market (19.4%). The difference between the contamination rate of the samples collected in the markets was not statistically significant (*p* < 0.05) (Table 4).

**Table 4:**
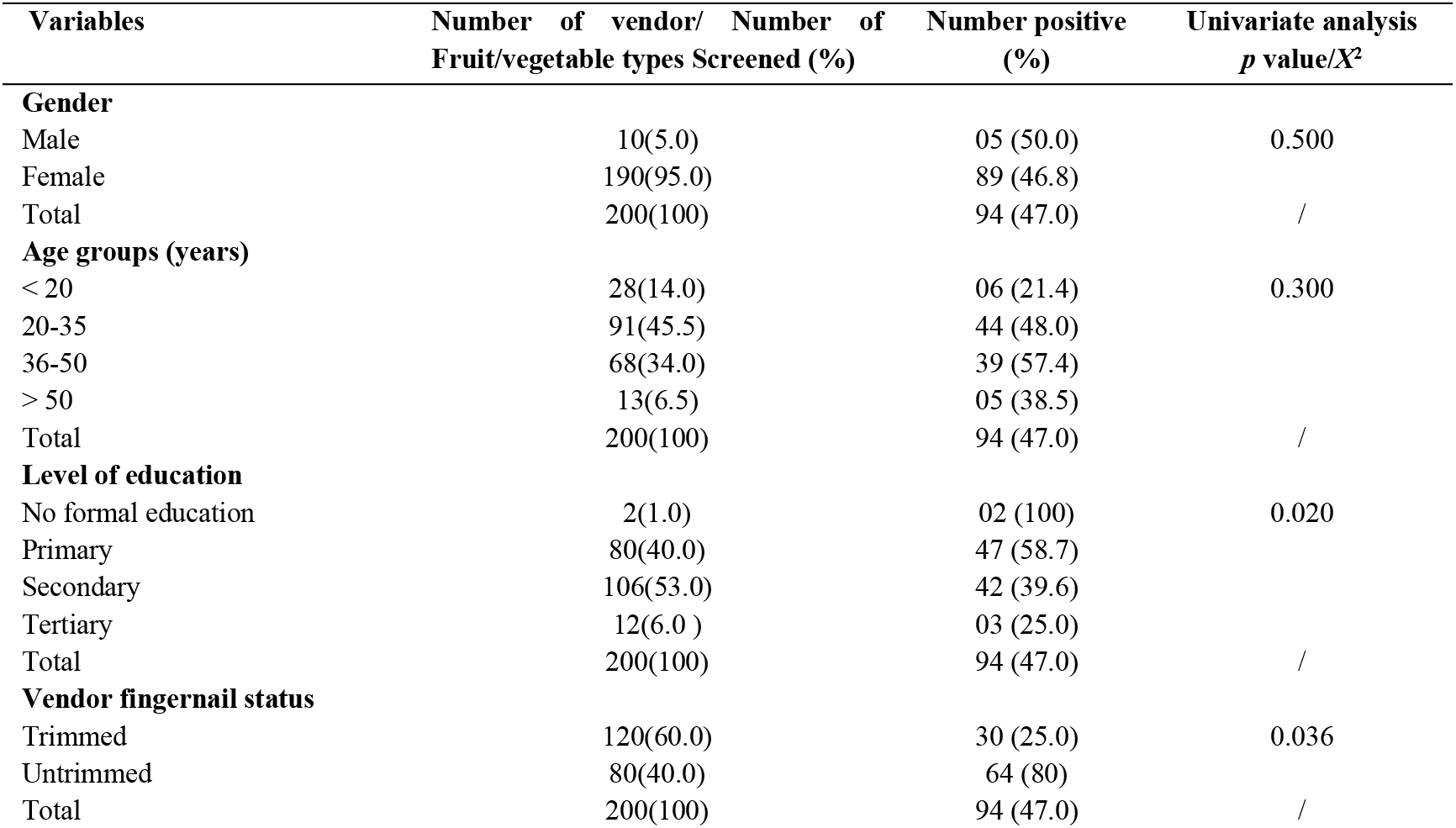
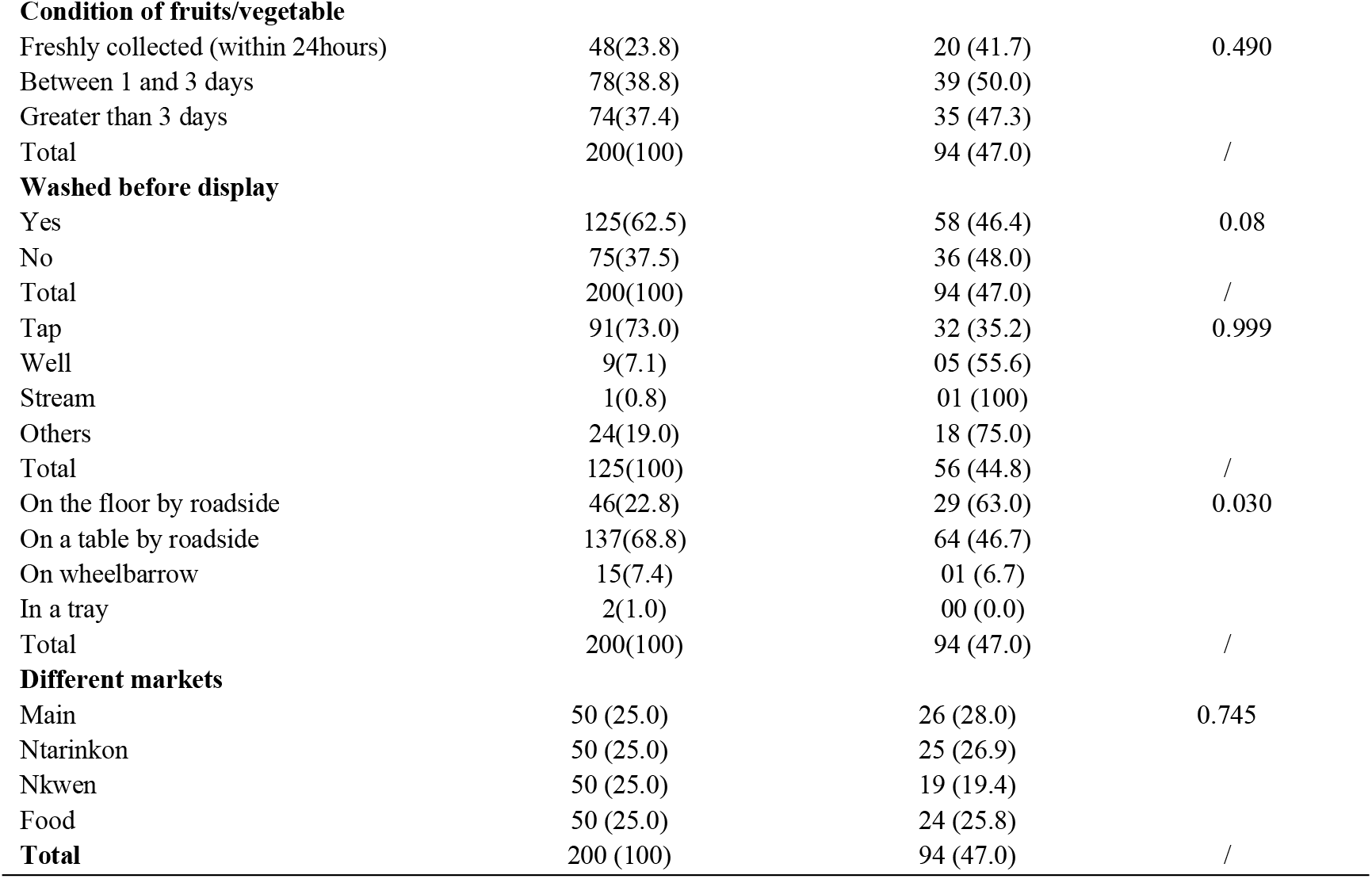
Univariate test of factors associated with parasitic contamination of fruits and vegetables sold in selected markets of Bamenda city.

## Discussion

The present study assessed parasitic contamination status and associated factors of fruits and vegetables in 4 different markets in Bamenda city. The overall parasitic contamination rate was found to be 47.0%, which is not far from 44.8% reported by Ebrahimzadeh *et al*. [10] in Iran, from 39.1% found in Ethiopia [29]. It is however higher than 14.6% reported in similar studies carried out by Rahmati *et al*. [30] in west of Iran and slightly lower than those obtained by Tefera *et al*. [28] (57.8%) in Ethiopia and Idahosa *et al*. [11] (56.25%) in Nigeria. The difference could be due to variations in items of samples collected, processing and laboratory methods used suggested by Alemu *et al*. [29]. One of the explanation for the high contamination rate in this study is that data for the present study was collected in the rainy season when the transmission, and hence the prevalence, is relatively high. The detection of intestinal parasites from fruits and vegetables is indicative of the fecal contamination from human and or animal origin. Isolation of more than one parasite per sample in this work reflects the possibility of a poly fecal contamination of vegetables that most probably result to poly parasitic infection in man. The high occurrence of these parasites reflects a high-level contamination and persistence of human infection. The discrepancy in the results of this study and previous studies may be due to geographical location, climatic, environmental conditions, the general behavioral attitude to hygiene and the socio-economic activities of producers, sellers and consumers. Equally, the kind of sample and sample size examined, the sampling techniques, methods used for detection of the intestinal parasites, and the seasonal period differs also, and consequently, the results differ variously. For instance, in the study from Egypt, samples were washed with tap water for 6-7 min for removal of mud and dust before being immersed in physiological saline [31]. This could decrease as some of the parasites might be removed with mud and dust particles. A study from Sudan screened only 150 samples and non-leafy items were screened and samples were not processed by modified acid-fast staining; therefore oocysts of intestinal coccidian were not assessed; all contributing to a lower contamination rate compared to the present study [32].

Carrot was the most frequently contaminated followed by cabbage and watermelon while pawpaw was the least to be contaminated. This probably due to the fact that produce with smooth surfaces provided minimum support for adhesion by parasites while produce like Carrot and Cabbages with uneven surfaces made the parasitic stages attach more easily to the surface. Vegetables are more prone to contamination than fruits due to the fact that vegetables like lettuce, spinach and cabbage have uneven/rough surfaces which enable parasites to attach more easily and overcome the effects of washing [26, 33]. Owing to the softness and fragility of leaves of vegetables, most vendors do not thoroughly wash them before display. On the contrary, the smooth surface of fruits like green pepper, tomato and mango might reduce the rate of parasitic attachment and can be washed easily. Edible parts of vegetables grow closer to the soil than that of fruits. Hence, soil may also play significant role in the contamination of vegetables [33, 34]. In support of this, vegetables were more likely to be contaminated as compared to fruits (*p* <0.05) in the present study. Use of human and animal excreta as an organic fertilizer might contribute for this contamination as confirmed by a couple of studies in Ethiopia [29]. *Cryptosporidium spp* and *Entamoeba hystolitica* were more frequent. Our finding concerning *E. hystolitica* (12.5%) was lower than reports from Sudan (42.9%) and Dessie town (24%), but higher than results from Arba Minch (8.4%)[36-38]. This might be due to the fact that the parasite have living state abundant in the environment hence easily contaminating fruits and vegetables as suggested by Tefera *et al*. [28], Alemu *et al*. [25]. Variation in the prevalence of *E. histolytica* might be attributed to the long periods of survival of the cysts under cool and moist conditions and variations in geographical distribution [37,38].

According to the present study, soil-transmitted helminthes (*A. lumbricoides* and hookworms) were predominant. Similar result was found in Nigeria and Arba Minch where the occurrence of *A. lumbricoides* and hookworms were predominant was 56.31% and 20,83% respectively. But our results disagree with findings in Oyo State of Nigeria where *A. lumbricoides* and hookworms were among the least detected parasites [30,39-42]. The difference of the result might be due to the effect of biannual deworming programme targeting soil transmitted helminthes and that is ongoing in those areas making the prevalence to drop as suggested by Alemu *et al*. [29].

Findings from multiple logistic regression analysis revealed that fruits and vegetables displayed without washing were more contaminated with parasites compared to those washed before display. The present findings are consistent with a study conducted in Dire Dawa, Arba Minch, and Jimma [5,15,21]. In fact, washing before display removes parasites. However, the actual finding disagree with the report by Bekele and Shumbej in Arba Minch [10,29]. The cleanliness of water used for washing and the washing process might bring such discrepancies. Also produce displayed in a bucket immersed in water appeared more contaminated than those sell on the shelf. This result may be probably due to the cross contamination during immersion.

Vendor with untrimmed fingernails were likely to collect dirt and parasite stage and contaminate produce touched. This finding explained why they had fruits and vegetables more contaminated than those with trimmed fingernails. In the present study 2.0% of parasites species were unidentified and it was not possible to separately report only on parasite of human medical importance since they were alive but morphologically indistinguishable.

The polyparasitic contamination observed our finding is similar with the result obtain by Alemu *et al*. [30], our result might indicate the possibility of high level contamination of the fruits and vegetables, which perhaps results in multiple parasitic infections in human. It might also indicate the persistence of intestinal parasitic infection in the area.

## CONCLUSION

The overall prevalence of parasitic contamination was 47.0%. Sixteen (16) different parasites were identified either in mono or in polyinfection. *Cryptosporidium pavum* and *Entamoeba histolytica* were protozoa more common, while *Ascaris lumbricoïdes* and Hookworms were the most frequent helminths. The act of washing the produces before displaying for sale, the sources of water used for washing, the means of display for selling and the duration of fruits and vegetables in the market appeared to be factors associated with parasitic contamination. These findings underscore the public health implication of fruit and vegetable farmers, sellers and consumers, being at high risk of infection with parasites.

## Data Availability

in Excel and SPSS softwares

## Acknowledgements

The authors thank the administrative staff of Bamenda Regional Hospital especially for providing Laboratory space for this research to be conducted. We also thank the vendors who accepted to participate in this study.

## Author contributions

KHLF and NNF conceived and designed the study, NHN collected the data, analysed, interpreted the results, KHLF, NNF and FO drafted and critically revised the manuscript. NNF and FO participated during the fieldwork and revised the manuscript. All authors read and approved the final manuscript.

